# Social experiences during participation in a community physical activity initiative: a cross-sectional study of parkrun participants in Ireland

**DOI:** 10.1101/2025.06.18.25329855

**Authors:** Allison Dunne, Alice Bullas, Helen Quirk, Steve Haake

## Abstract

Community initiatives are a popular social prescribing choice to support mental wellbeing. Recent studies suggest that parkrun, a global physical activity initiative, can promote mental wellbeing and social aspects may be a contributing factor. This study considers the relationship between parkrun participation (as a runner/walker or volunteer) and the social interactions and opportunities it provides. Differences in the social experience with regards to parkrun participation type, gender, and age are explored. A secondary data analysis of a cross-sectional survey of 4,304 parkrun participants in Ireland showed that 30.4% of respondents had high mental wellbeing scores. The most common social interaction was with *other parkrun participants* at 69.4%, and 48.2% said *I feel part of my community* at parkrun. People who volunteered at parkrun (in addition to running/walking) were more likely to report social relationships and opportunities than those who did not volunteer. Females were more likely than males to feel isolated at parkrun (20.9% vs 15.4%, p<0.001). The results suggest that parkrun participation may provide a variety of social experiences which supports the inclusion of parkrun in the suite of social prescribing offerings. Providing a supportive environment for social interaction could be a health promotion recommendation for physical activity interventions to enhance the mental wellbeing impacts.

## Background

Maintaining good mental wellbeing is important for quality of life and is a key protective factor against future mental health conditions (Barry, Clarke, Petersen, & Jenkins, 2019; Keyes, Dhingra, & Simoes, 2010). Influences on mental wellbeing include lifestyle, social and economic circumstances, as described in Dahlgren and Whitehead’s social determinants of health model (Dahlgren & Whitehead, 2021). Living in areas of deprivation (Sorin et al., 2022) or in rural communities, where isolation and loneliness are common can lead to poor mental wellbeing, particularly for men in Ireland (O’Donnell & Richardson, 2020). Poor mental wellbeing is a risk factor for suicide (Department of Health, 2015). Data from deaths by probable suicide in Ireland from 2015 to 2018 (Cox, Munnelly, Rochford, & Kavalidou, 2022) show the highest numbers of probable suicides were from the age group 35-54 years (all genders) and 76% of probable suicides from all age groups were by men; the age group with the highest number of deaths for men was 40-44 years (Cox et al., 2022).

Social and community networks are influential on mental wellbeing since a support system can help to build resilience against mental health challenges (Costello, McDermott, Patel, & Dare, 2019). Sense of belonging and social cohesion provided through meaningful social activities benefit mental wellbeing (Forsman, Herberts, Nyqvist, Wahlbeck, & Schierenbeck, 2013; Williams, Maguire, Morrissey, Taylor, & Wyatt, 2020). Even brief interactions with strangers have been shown to positively impact happiness and wellbeing (Sandstrom & Boothby, 2021; Van Lange & Columbus, 2021). This has been recognised in recent policies such as Sharing the Vision, the national mental health policy for the Republic of Ireland (referred to as Ireland in the remainder of this paper), which highlights the importance of social support and community groups in the management of pre-clinical mental health challenges (Department of Health, 2020). The policy includes social prescribing, a formal referral system to link patients to community initiatives, to protect mental wellbeing (Department of Health, 2020; Health Service Executive, 2021). These initiatives often include a physical activity element (for example; walking, gardening or swimming) along with an opportunity for social interaction (Chatterjee, Camic, Lockyer, & Thomson, 2018; Costello et al., 2019; Leavell et al., 2019). The Irish National Sports Policy recognises that participation in physical activity and associated volunteering roles are important for community development and social cohesion (Government of Ireland, 2018). One community physical activity initiative which has a growing body of research in relation to social prescribing is parkrun.

parkrun (stylised with a small p), is a free, weekly, 5 km running, walking, and volunteering community initiative organised with the support of the international parkrun charity. Starting in the UK in 2004, parkrun events are now held in 23 countries worldwide (parkrun, 2024). The first parkrun in Ireland took place in Malahide, Dublin, in 2012 (parkrun, 2020). Ireland currently has parkruns on Saturday mornings in more than 100 locations with over 400,000 people registered (Shields, 2023) and weekly attendance of over 11,000 runners or walkers and 1,500 volunteers (based on figures on 24/08/24) (parkrun Ireland, 2024).

Although parkrun events centre around physical activity there is an important social element. In addition to social contact before, during, and after participation (Morris & Scott, 2019), many locations offer participants the opportunity to socialise in a café after the event (Bowman, 2018). Studies from the UK and Australia suggest that social support and social capital can be used to initiate and maintain parkrun attendance (Chivunze, Burgess, Carson, & Buchholtz, 2021; Sharman, Nash, & Cleland, 2019; Wiltshire & Stevinson, 2018). A study of UK parkrunners who were inactive before commencing parkrun (Wiltshire & Stevinson, 2018) explores how social capital can be used to initiate and maintain parkrun participation. The study noted exchange of knowledge and support of new parkrunners, highlighting that social support can be accessed through developing social capital (Wiltshire & Stevinson, 2018, p. 55). Social support was described by one participant in the context of helping a subjectively unfit runner to complete the event (Wiltshire, Fullagar, & Stevinson, 2018).Bowness, Tulle and McKendrick (2020) describe parkrun as a “sacred space” for communities which can strengthen social bonds within and beyond families, explaining that the feeling of social connection was what differentiated parkrun participation from running or walking alone. Social connections have also been demonstrated in parkrunners from the UK with mental health conditions (Morris & Scott, 2019) with volunteering facilitating additional opportunities for social interaction in this population (Ashdown-Franks et al., 2023).

The aim of this study is to explore whether the social interactions and opportunities at parkrun are the type which have potential for supporting mental wellbeing in an Irish population by answering the following questions:

1. Does parkrun participation as a runner, walker or volunteer provide a supportive environment for social interactions and social opportunities?
2. Is there a difference in the social experience of parkrun attendees with regards to participation type, gender, and age?

## Method

This study is a secondary analysis of data from a cross-sectional observational survey of parkrun Ireland registrants (the parkrun Health and Wellbeing Survey). Ethical approval was granted by Sheffield Hallam University Research Ethics Committee on 24/7/2018 for the parkrun Health and Wellbeing Survey (reference number: ER7034346) and the secondary analysis on 23/6/2020 (ER23166109). Both the original survey and this secondary data analysis were supported by the parkrun Research Board. The reporting of this manuscript adheres to established standards for reporting internet-based surveys using The Checklist for Reporting Results of Internet E-Surveys (CHERRIES) (Eysenbach, 2004).

### The parkrun Health and Wellbeing survey

The parkrun Ireland Health and Wellbeing Survey was hosted on Qualtrics (www.qualtrics.com) and distributed in English by email between 29th October 2018 and 3rd December 2018 to 189,550 adults in Ireland aged 16 and over who were registered with parkrun (Haake, Bullas, & Quirk, 2019). Of 8,612 returned surveys (4.5% response rate), 4,304 respondents completed the questions (2.3% completion rate). The remaining 4,308 either opened the survey and did not consent to participate (153), opened the survey and consented to participate but did not answer any questions (3,725), were duplicates (189) or completed the survey and self-identified as *registered but not yet participated* (241). Those *registered by not yet participated* fell out of the remit of this study and are not included in the analysis.

A comprehensive outline of the survey design, content and methods is contained within reports from Haake et al. and Quirk et al. (Haake et al., 2019; Quirk et al., 2021). A full copy of the parkrun Health and Wellbeing Survey can be found in Haake et al (2019). Briefly, the survey asked 47 questions relating to various aspects of mental and physical health and wellbeing, alongside self-reported participation type: volunteers, runners/walkers who volunteer, runners/walkers (who do not volunteer).

Where respondents recorded their parkrun identification number, parkrun participation data was matched to survey responses. At the time of the survey, the only options for gender identity at parkrun registration were *male*, *female* or *prefer not to say*. This has since been updated to include a fourth option of *another gender identity.* Data extracted from the full Health and Wellbeing Survey dataset consisted of the responses to questions on mental wellbeing, social interaction, and social opportunities, along with demographic information. Mental wellbeing was measured using the Short Warwick Edinburgh Mental Wellbeing Scale (SWEMWBS) (Warwick Medical School, 2021).

The questions on social interactions and social opportunities were developed for the Health and Wellbeing Survey and presented as follows:

> Social interactions: “Who do you usually interact with at parkrun? i.e. those you attend and communicate with at parkrun.” A choice of 10 options was provided (*family, friends, parkrun participants, strangers, spouse/partner, member of a sports club, member of a non-sports club, neighbours, work colleagues and did not interact)*, with survey respondents free to select any number of options that applied without restriction. A chance to describe *other* using free text was included.

> Social opportunities: “In terms of relationships, what opportunities has parkrun opened up for you?” A choice of 8 options was provided (*I have met new people of a similar background, I have met new people of a different background, I feel closer to my existing friends or family, I have joined a sports club, I have joined a non-sports club, I feel part of my community, it’s made me feel isolated, it has allowed me to spend time on my own*) with the survey respondents free to select any number of options that applied without restriction. A chance to describe *other* using free text was included.

Respondents were segmented into three age categories (16-44 years, 45-64 years and 65+ years), taking into consideration target ages for suicide prevention and also chosen to allow comparison with previous research such as Grunseit et al (2017)

### Data preparation

Data validation and preparation for statistical analysis were completed using Excel for PC (V16, Microsoft Corporation). The initial handling and validation of raw data from the survey are described in a previous publication (Quirk et al., 2021). As some survey respondents did not answer all questions, data tables include the relevant sample size (n).

### Statistical analysis

Statistical analysis was used to compare results within the three demographic groups; gender, age, and parkrun participation type. The chi-square test of independence (χ2) statistic was used to evaluate the differences between social and mental outcomes because two of the three groups, gender and parkrun participation type, had categorical data. The age groups are ordinal data, so more detail could be obtained by using a different statistical test, such as Mann-Whitney U test, but to allow for a consistent set of results it was decided to use χ2 for all three groups of data. Effect size less than 0.1 was considered small, 0.3 medium, and 0.5 large (Field, 2018). Cross tabulation and statistical analysis were conducted using IBM SPSS Statistics for PC (v26). Statistical significance *α* level was set as *p*<0.01.

Bivariate correlations were used to look for associations between the different social connections and social opportunities. A stepwise multiple linear regression was carried out with the SWEMWBS score as the response variable and gender, age (in years), social connections and social opportunities as the predictor variables with a selection threshold for each variable set at *p*<0.05.

## Results

Data were analysed from 4,304 respondents to the parkrun Health and Wellbeing Survey Ireland (Haake et al., 2019). Table 1 shows that 54.4% of respondents were female (compared to 56.6% of the whole parkrun population at the time of the survey) and were predominantly white (98.5%). At the time of writing, ethnicity data was not collected at the time of parkrun registration, so the data cannot be compared. The scores for the Short Warwick Edinburgh Mental Wellbeing Scale (SWEMWBS) showed a skew towards high mental wellbeing levels with 30.4% of survey respondents recording high scores (Table 1).

**Table 1.** Characteristics of survey respondents.

|  | n | % |
| --- | --- | --- |
| All respondents | 4,304 |  |
| **Gender (n=3,413)** |  |  |
| Male | 1,558 | 45.6 |
| Female | 1,855 | 54.4 |
| **Age category in years (n=4,303)** |  |  |
| 16-44 | 1,990 | 46.2 |
| 45-64 | 2,103 | 48.9 |
| 65+ | 210 | 4.9 |
| **parkrun participant type (n=4,304)** |  |  |
| Runner or walker and volunteer | 1,967 | 45.7 |
| Runner or walker | 2,283 | 53.0 |
| Volunteer | 54 | 1.3 |
| **Ethnicity (n=4,270)** |  |  |
| White Irish | 3,794 | 88.9 |
| White Irish Traveller | 3 | 0.1 |
| White - other | 406 | 9.5 |
| Black, Asian, other minority | 50 | 1.2 |
| Rather not say | 17 | 0.4 |
| **Mental wellbeing SWEMWBS (n=4,304)** |  |  |
| Low (7-19) | 409 | 9.5 |
| Average (20-27) | 2,586 | 60.1 |
| High (28-35) | 1,309 | 30.4 |

### Social interactions

Responses to the question “Who do you usually interact with at parkrun?” are shown in Figure 1 (with data in Tables S1, S2 and S3 in supplementary material) segmented by gender (Figure 1a), age (Figure 1b) and participation type (Figure 1c). The most selected response for social interaction was other *parkrun participants* (69.4%). Interactions with *friends* and *strangers* showed similar levels of response (45.9% and 44.1% respectively), while 5.9% noted that they *did not interact* with anyone at parkrun. There were no significant differences in responses between males and females for any response (Figure 1a).

**Figure 1a, 1b, 1c.**
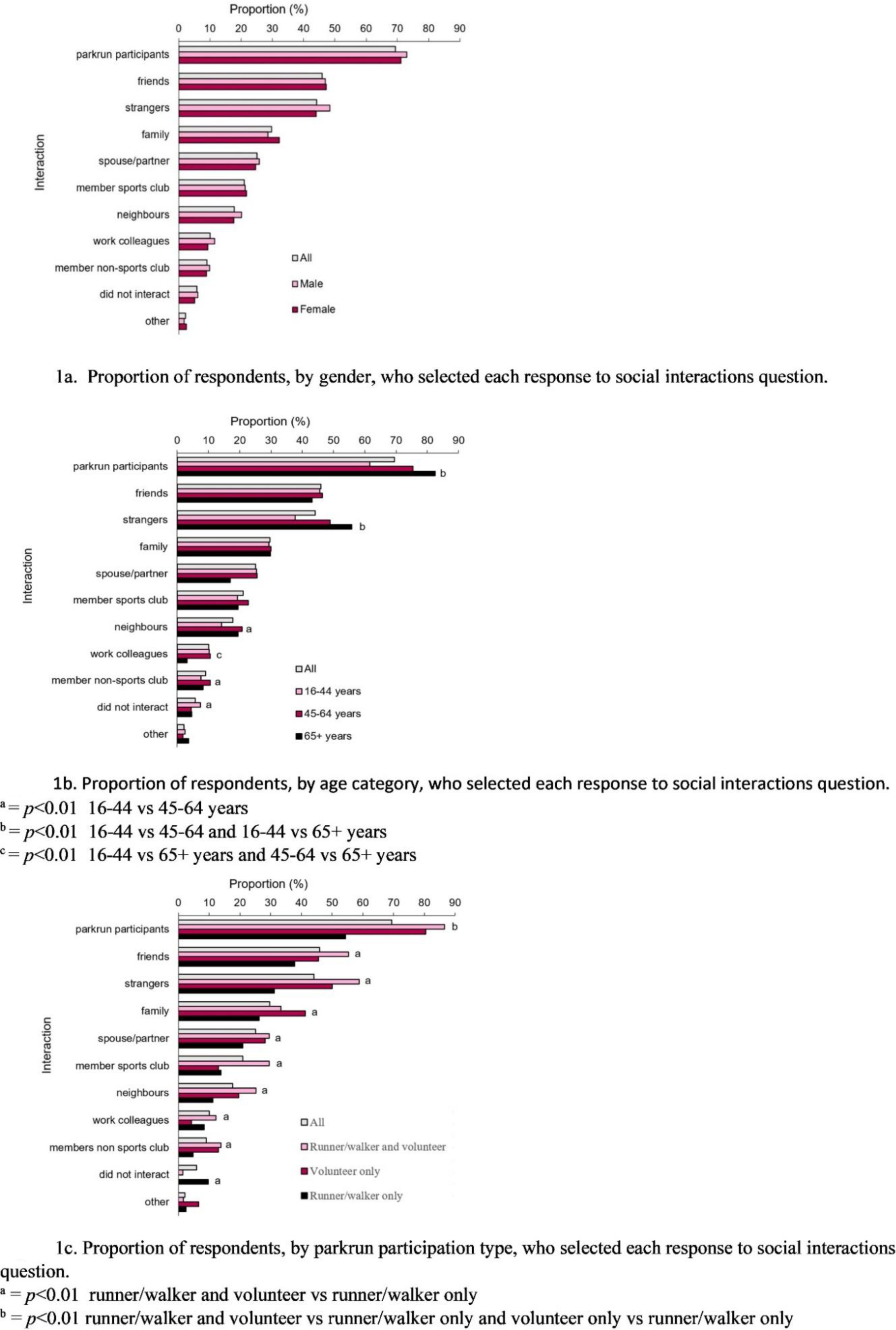
Proportion of respondents who selected each response to social interactions question.

Figure 1b shows that younger participants (age 16-44 years) were least likely to note social interactions, when compared to participants over 45 years old. When considering the responses for each type of interaction the youngest age group were less likely (than 45-64 years and over 65 years) to interact with other *parkrun participants* and *strangers* (both *p*<0.001). There were no age differences for interacting with *friends, family, spouse, or member of a sports club*.

Responses segmented by the parkrun participation type are shown in Figure 1c. Respondents who volunteered at parkrun, both with or without running/walking, were significantly more likely to interact with *other parkrun participants* (*p*<0.001). For other choices, runners/walkers who volunteer were often the most likely to note a social interaction; for example, with *friends, strangers*, *spouse/partner*, member of a club (*sports* and *non-sports club*) and *work colleagues* (all *p*<0.001). Respondents who volunteered without running/walking were most likely to note an interaction with *family* (*p*<0.001). Runners/walkers (who did not volunteer) were most likely to say that they *did not interact* with anyone at parkrun (*p*<0.001).

Table 2 shows bivariate correlations between the social interaction questions with moderately-sized correlations in bold (r>0.2). It can be seen the selection of *parkrun participants* was moderately correlated with the selection of *strangers* and inversely correlated with the choice of *did not interact*. The selection of *neighbours* was also moderately correlated with the selection of *friends*, while the selection of *members of sports clubs* was moderately correlated with the choice of *members of non-sports clubs*. The choice of *family* was moderately correlated with the choice of *spouse/partner*.

**Table 2.**
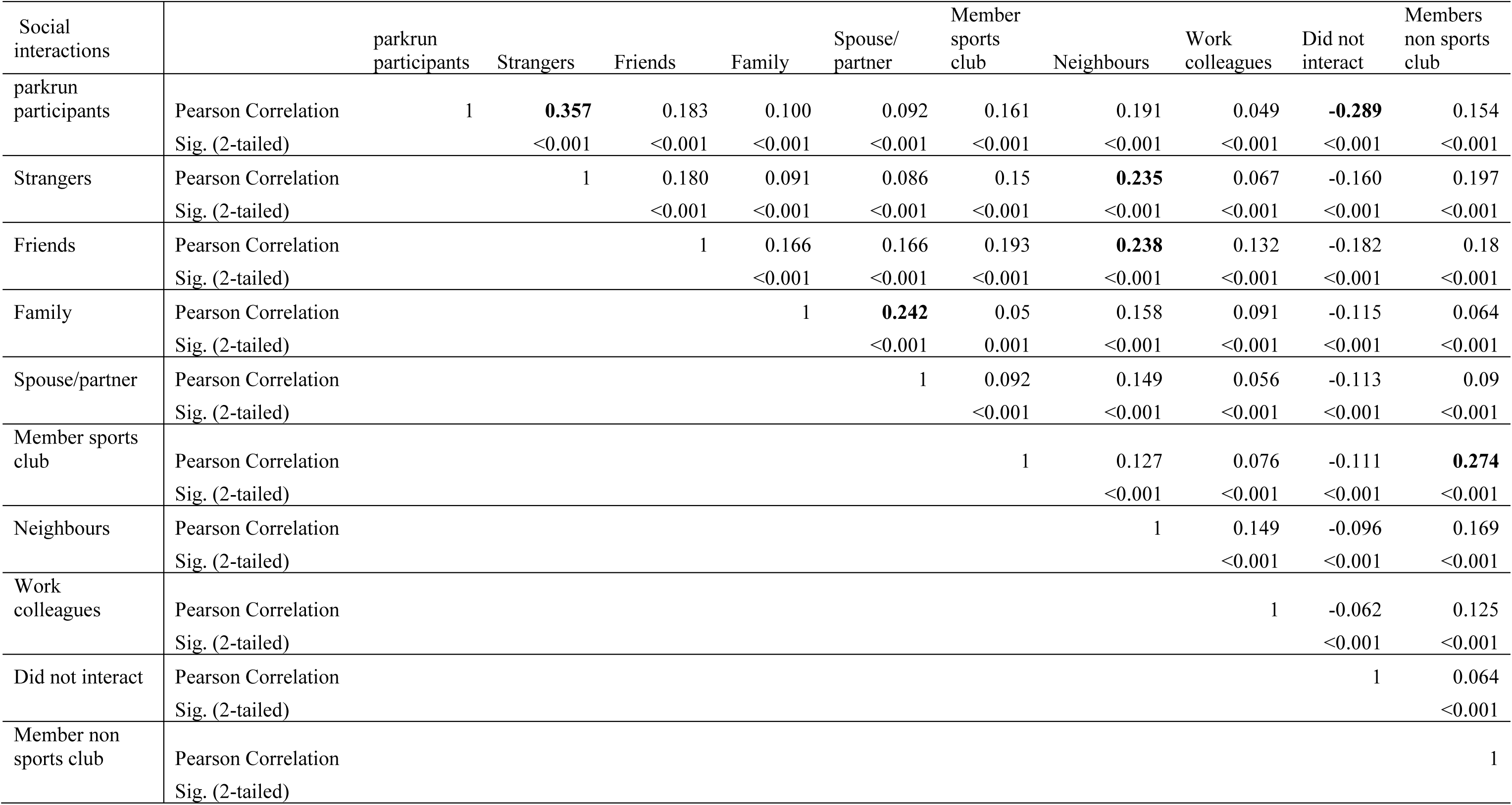
Bivariate correlation for social interactions.

### Social opportunities

Responses to the question “In terms of relationships, what opportunities has parkrun opened up for you?” are shown in Figure 2 (with data in Tables S1, S2 and S3 in supplementary material) segmented by gender (Figure 2a), age (Figure 2b) and participation type (Figure 2c). The most common responses were *I feel part of my community* (48.2%), *I have met new people of a different background* (33.7%) and *I have met new people of a similar background* (32.4%). The response *It has made me feel isolated* was chosen by 18.5% of respondents. Two response types showed a significant gender difference (Figure 2a): females were more likely to select *It’s made me feel isolated* (*p*<0.001) but less likely than males to select *It’s made no difference to me* (*p*=0.004).

**Figure 2a, 2b, 2c.**
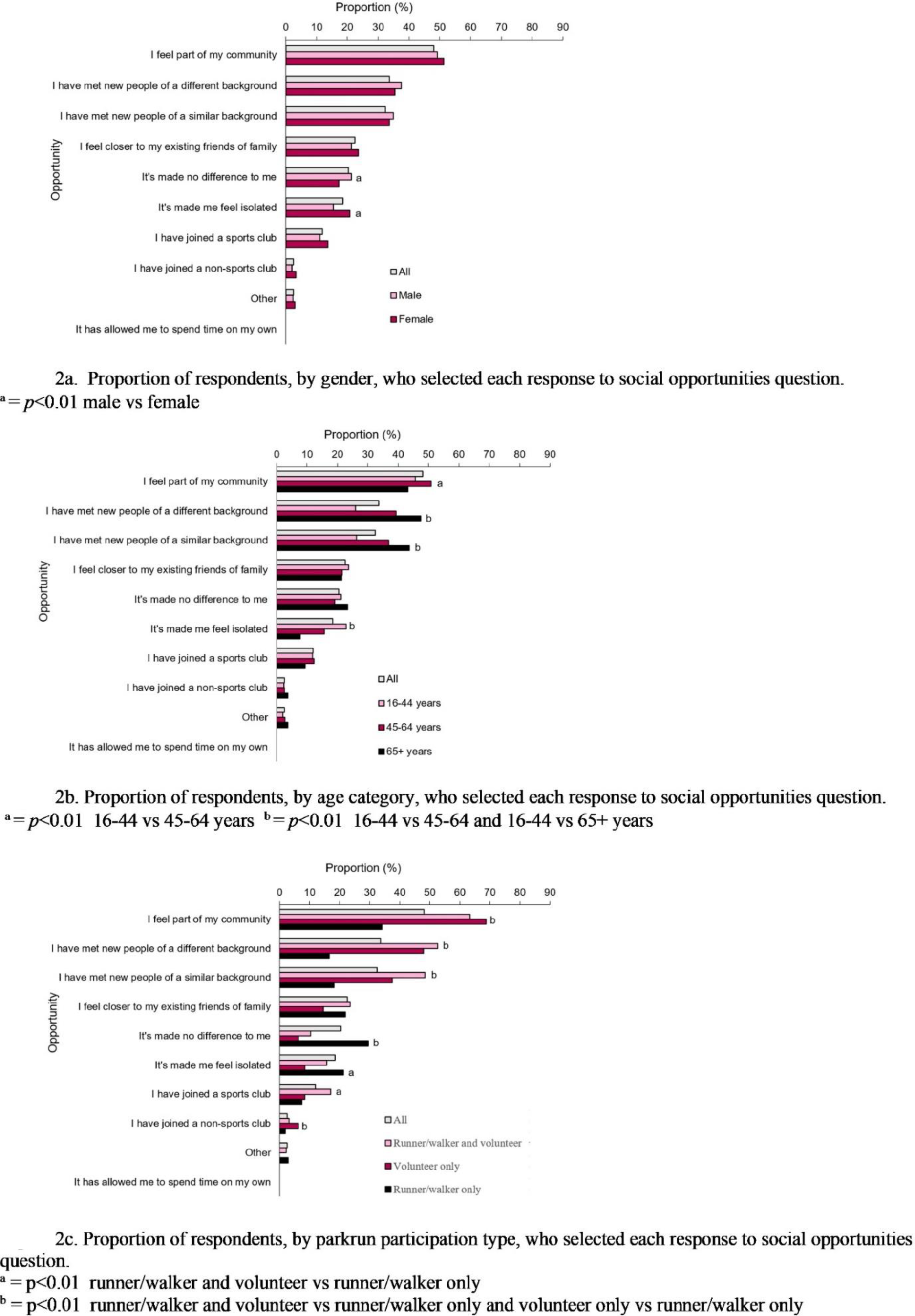
Proportion of respondents who selected each response to social opportunities question.

There were some age group differences in the responses to the social opportunities question (Figure 2b): 45-64 year olds were most likely, when compared to the other two age groups, to say they *felt part of the community* (*p*=0.002). Over 65 year olds were most likely to select *I have met new people*, from both *similar* or *different backgrounds* (*p*<0.001) while the youngest respondents (16-44 years) were most likely to say that parkrun participation *made me feel isolated* (*p*=<0.001).

There were several differences in responses to social opportunities between the parkrun participation types with respondents who volunteer, with or without running/walking, more likely to note social opportunities (Figure 2c). Respondents who volunteer without running/walking were most likely to say they *felt part of my community* (*p*<0.001). Runners/walkers who volunteer were most likely to have *met new people*, both from a *similar* or *different background* (*p*<0.001). Runners/walkers (who don’t volunteer) were most likely to state *It’s made no difference to me* and *It’s made me feel isolated* (*p*<0.001).

Table 3 shows bivariate correlations between the social opportunity questions, again with moderately-sized correlations in bold (r>0.2). The choice *I feel part of my community* was moderately correlated with the choice *I have met people of a different background* and *I have met new people of a similar background*, and inversely correlated with *it’s made no difference to me*.

**Table 3.**
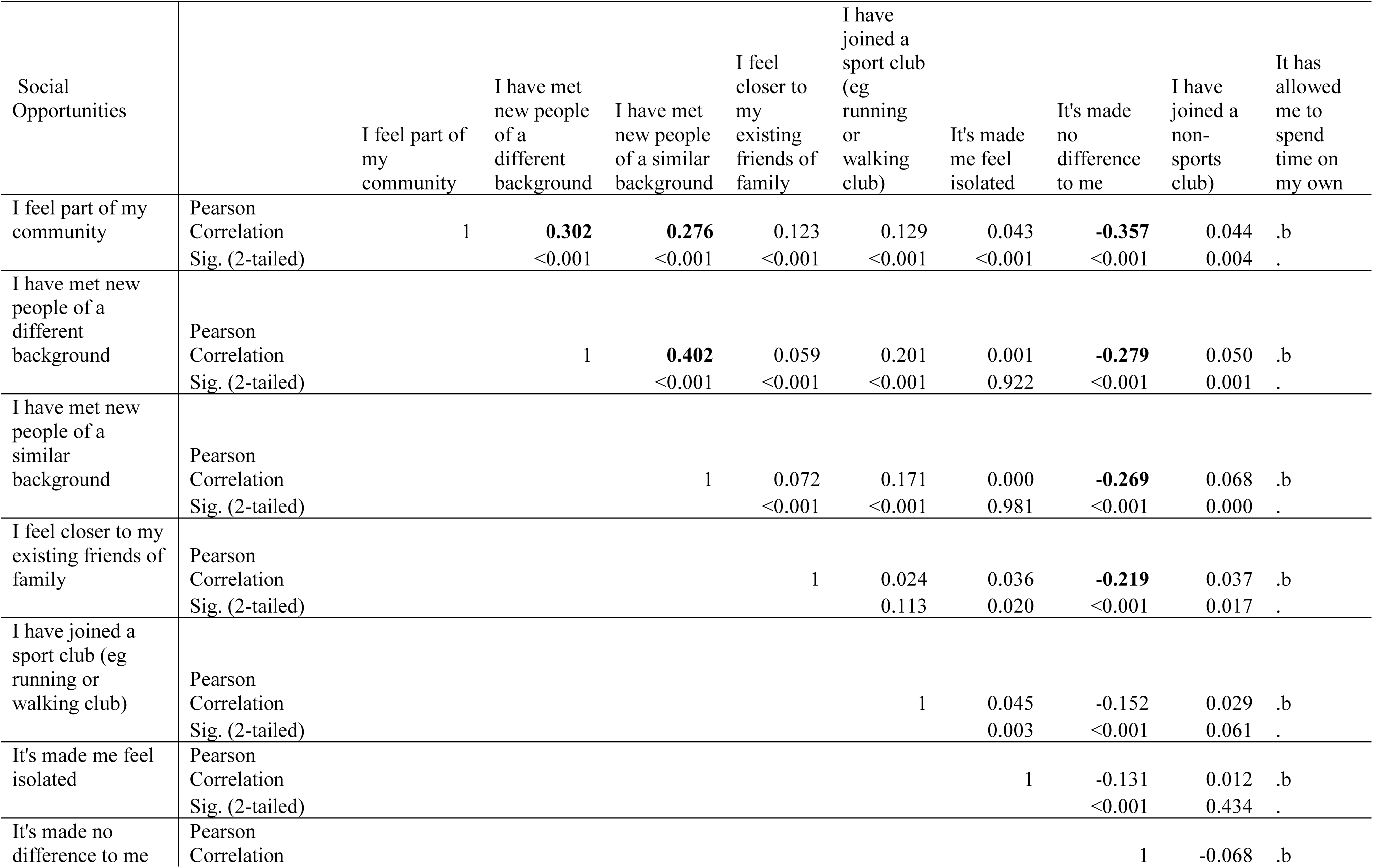
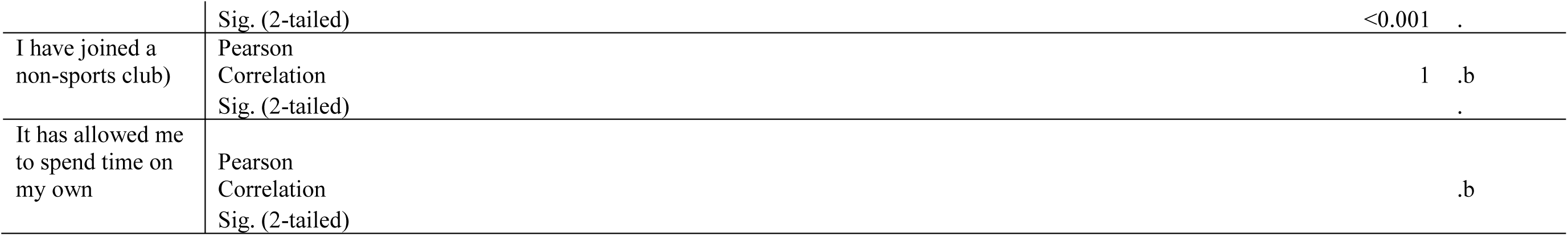
Bivariate correlation for social opportunities.

| Social Opportunities |  | I feel part of my community | I have met new people of a different background | I have met new people of a similar background | I feel closer to my existing friends of family | I have joined a sport club (eg running or walking club) | It's made me feel isolated | It's made no difference to me | I have joined a non-sports club) | It has allowed me to spend time on my own |
| --- | --- | --- | --- | --- | --- | --- | --- | --- | --- | --- |
| I feel part of my community | Pearson Correlation<br>Sig. (2-tailed) | 1 | **0.302**<br><0.001 | **0.276**<br><0.001 | 0.123<br><0.001 | 0.129<br><0.001 | 0.043<br><0.001 | **-0.357**<br><0.001 | 0.044<br>0.004 | .b<br>. |
| I have met new people of a different background | Pearson Correlation<br>Sig. (2-tailed) |  | 1 | **0.402**<br><0.001 | 0.059<br><0.001 | 0.201<br><0.001 | 0.001<br>0.922 | **-0.279**<br><0.001 | 0.050<br>0.001 | .b<br>. |
| I have met new people of a similar background | Pearson Correlation<br>Sig. (2-tailed) |  |  | 1 | 0.072<br><0.001 | 0.171<br><0.001 | 0.000<br>0.981 | **-0.269**<br><0.001 | 0.068<br>0.000 | .b<br>. |
| I feel closer to my existing friends of family | Pearson Correlation<br>Sig. (2-tailed) |  |  |  | 1 | 0.024<br>0.113 | 0.036<br>0.020 | **-0.219**<br><0.001 | 0.037<br>0.017 | .b<br>. |
| I have joined a sport club (eg running or walking club) | Pearson Correlation<br>Sig. (2-tailed) |  |  |  |  | 1 | 0.045<br>0.003 | -0.152<br><0.001 | 0.029<br>0.061 | .b<br>. |
| It's made me feel isolated | Pearson Correlation<br>Sig. (2-tailed) |  |  |  |  |  | 1 | -0.131<br><0.001 | 0.012<br>0.434 | .b<br>. |
| It's made no difference to me | Pearson Correlation |  |  |  |  |  |  | 1 | -0.068 | .b |

|  |  |  |  |
| --- | --- | --- | --- |
|  | Sig. (2-tailed) | <0.001 | . |
| I have joined a non-sports club) | Pearson Correlation | 1 | .b |
|  | Sig. (2-tailed) |  | . |
| It has allowed me to spend time on my own | Pearson Correlation |  | .b |
|  | Sig. (2-tailed) |  |  |

### Mental wellbeing score relationship with social connections and opportunities

Table 4 shows the results of a stepwise linear regression for the Short Warwick Edinburgh Mental Wellbeing Scale score. For a given age, the model estimates the impact of selecting each connection or opportunity on mental wellbeing score. The overall regression was weakly significant (R^2^ = 0.066, F(6, 3151) = 37.514, p < 0.001).

**Table 4.** Coefficients from stepwise multiple linear regression for Short Warwick Edinburgh Mental Wellbeing Scale Score.

|  | Unstandardised Coefficients |  | Standardised Coefficient | t | Sig. |
| --- | --- | --- | --- | --- | --- |
|  | B | Std. Error | Beta |  |  |
| (Constant) | 21.243 | 0.308 |  | 69.055 | <0.001 |
| Age (years) | 0.076 | 0.006 | 0.218 | 12.561 | <0.001 |
| Friends | 0.452 | 0.147 | 0.055 | 3.079 | 0.002 |
| Family | 0.436 | 0.156 | 0.049 | 2.791 | 0.005 |
| I feel part of my community | 0.434 | 0.145 | 0.053 | 2.984 | 0.003 |
| It's made me feel isolated | -0.574 | 0.187 | -0.053 | -3.07 | 0.002 |
| Work colleagues | 0.470 | 0.235 | 0.035 | 1.996 | 0.046 |

The fitted regression model was, Mental wellbeing score (SWEMWBS)= 21.711+0.076(Age)+0.452(Friends)+0.436(Family) +0.470(Work colleagues)+0.434(I feel part of my community)-0.574(It’s made me feel isolated) where age was in years, and values for social connections and opportunities were 0 when not selected and 1 when selected. The standard error of the estimate was 4.0.

## Discussion

The results of this study support the idea that parkrun is more than just a physical activity initiative. The combination of an activity for all ages with volunteering as an option allows for social interactions and opportunities.

The results showed clear differences across the age categories. Participants over 45 years were more likely to interact with a wide range of people at parkrun and noted more relationship opportunities than younger participants. Previous research proposed that younger parkrunners are focussed on the physical health benefits of parkrun rather than psychological wellbeing (Grunseit et al., 2017). The current study echoes these results and suggests that older people could view parkrun as a social or community initiative rather than purely a physical challenge. This makes parkrun a suitable recommendation, via social prescribing, for the over 45 age group to increase their social network.

parkrun participation being linked to community connection was shown to be very strong among survey respondents. Similar results were shown by Grunseit et al. (2017) where parkrun participants age 50-64 years had the highest scores for community factors. Further exploration of the mechanism for the community connection and how it impacts people of different ages would be insightful.

The social experiences provided by parkrun suggest that people have the opportunity to spend time with their existing social circles (e.g., friends/family members) but also people who they consider ‘different’ to themselves. The social opportunities statements *I have met new people of a similar background* and *I feel closer to my existing friends and family* are indicative of bonding social capital. Social capital relates to the worth of interpersonal relationships taking into account the size of network of social connections and how much value each connection has (Bourdieu, 1986). Putnam developed this concept by proposing two forms of social capital: bonding and bridging (Helliwell & Putnam, 2004; Putnam, Leonardi, & Nanetti, 1993). Bonding capital describes connections between individuals whereas bridging capital explores relationships beyond this network to “bridge” to individuals or organisations outside the usual social circle to provide benefits to the people in that network (Ferlander, 2007; Helliwell & Putnam, 2004). The concept of linking capital is an extension of bridging capital which describes relationships between people with different levels of power or authority (Szreter & Woolcock, 2004).

Over 65s and the survey respondents who participated at parkrun as runners/walkers who volunteer. were the most likely groups to note social opportunities relating to bonding capital. This finding is particularly relevant for older participants who are at high risk of loneliness and social isolation (Dayson, Harris, & Woodward, 2021; Lindsay Smith, Banting, Eime, O’Sullivan, & van Uffelen, 2017). Participation in parkrun with the aim of increasing bonding social capital to support wellbeing would be a suitable social prescribing offering for this age group.

Over a third of respondents noted that they had met people from a different background to their own. This response was greatest for runners/walkers who volunteer and the over 65 age group. This is a positive sign in relation to building social capital as previous research suggests that parkrun attracts people who already know other people at parkrun (Sharman et al., 2019; Wiltshire & Stevinson, 2018). The results of this study show that by interacting with people from *different* backgrounds, parkrun provides an opportunity for the development of relationships outside of the usual social circle, a feature of bridging capital. The question did not ask any detail about the different background so it is impossible to differentiate from this study whether it was bridging or linking capital that was being impacted. Future studies could continue to explore the impact of parkrun participation on these types of social capital as interventions which promote both types of social capital are thought to be influential on population health (Kim, Subramanian, & Kawachi, 2006).

Correlations showed that the choice of one social connection was often associated with the choice of other, related social connections: for instance, *family* and *spouse/partner*; *friends* and *neighbours*; *members of sports clubs* and *non-sports clubs.* A similar effect was seen in the choice of opportunities in which those choosing *I feel part of a community* were associated with those choosing *I have met people of a similar* or *a different background*. Some of these selections were negatively associated with *did not interact* and *It’s made no difference to me*.

The multiple linear regression for mental wellbeing score controlled for some of these interactions and predicted that those identifying connections with friends, family and work colleagues, and feeling more part of a community were likely to have a higher mental wellbeing score. Those who felt isolated had a lower mental wellbeing score. The mental wellbeing score increased with age by around 0.74 per decade of age increase with associated increases in social connections and opportunities shown in Figures 1a, 1b and 1c and Figures 2a, 2b and 2c. The implication for parkrun is that attention might focus on interventions to increase social interaction for younger participants, especially those who don’t interact with others, or feel isolated.

Volunteering at parkrun, either as an activity in its own right, or in addition to running or walking, appeared to be associated with an increased likelihood social interactions and opportunities among the survey respondents. As the relationship could be bidirectional it could be the case that people who are naturally more confident and sociable are more likely to sign up for volunteering (Sandstrom & Boothby, 2021; Stride, Fitzgerald, Rankin-Wright, & Barnes, 2020). However, it also suggests that volunteering at parkrun plays a role in the social element of parkrun participation. Volunteering at parkrun could be linked to the concepts of reciprocity and civic engagement described by Putnam et al (1993) which has a role in building social capital and could explain why the survey respondents who volunteer at parkrun (with or without running/walking) noted a wider range of social relationships and opportunities (Hallett, Gombert, & Hurley, 2020). The theme of reciprocity was also mentioned by Stevinson, Wiltshire and Hickson (2015) where it was highlighted as a reason for initial and ongoing attendance at parkrun.

It would be useful to explore the depth of social interaction further as even quick social encounters, described as ‘casual sociability’ at parkrun by Hindley (2020), are potentially valuable for increasing social capital and have been shown to have positive effects on mood and sense of belonging (Sandstrom & Dunn, 2014). An exploration of the impact of casual encounters with strangers at parkrun or the building of stronger social networks will give a deeper understanding of the wellbeing benefits of parkrun in Ireland and across the parkrun portfolio globally.

The results of this study indicate that parkrun is an example of the type of social intervention recommended by Burgess, Jain, Petersen and Lund (2020) in their call to action for global mental health as it may have booster effects within the community to protect and enhance mental wellbeing. Mental wellbeing of the survey respondents was measured using the SWEMWBS tool. There are currently no published reference scores from the SWEMWBS for the Irish population. Comparing the scores from this study with a large population study in neighbouring country England indicates that the parkrun survey respondents from Ireland had a smaller proportion with poor mental wellbeing and greater proportion with high mental wellbeing scores (Ng Fat, Scholes, Boniface, Mindell, & Stewart-Brown, 2017). Although the mental wellbeing scores for this study were higher than we would expect in the general population, it is not possible to say from this cross-sectional study that parkrun participation has caused this increase. It could be that people with higher mental wellbeing are more motivated to attend parkrun. This is an area for further exploration, particularly as a similar phenomenon appears to be shown in Irish attendees of the Men’s Shed social initiative (McGrath, Murphy, Egan, & Richardson, 2022).

Not every group of survey respondents noted increases in social connections through parkrun participation. Runners/walkers who did not volunteer were most likely to note the feeling of isolation at parkrun. This supports the theory that volunteering at parkrun increases the sociability of the event by reducing the feeling of isolation and echoes the results from Ashdown-Franks et al (2023) who found that volunteering enhanced social inclusion at parkrun for people with mental health conditions. In the current study over 20% of female respondents and those aged 16-44 years of both genders said that parkrun made them feel isolated. Isolation is the opposite to social connectedness, is linked to loneliness, and could be a contributor to poor mental wellbeing (Dayson et al., 2021). There have been reports of perceived cliques at parkrun events which lead to a less inclusive atmosphere (Bowness et al., 2020; Fullagar et al., 2019). Cliques are an intense form of social network where members of the clique have strong ties but people outside of that group feel excluded (Chang, 2009) and can be considered as a form of bonding capital which leads to negative outcomes in society (Bourdieu, 1986; Nicholson & Hoye, 2008). It would be worthwhile investigating the mechanism for females and young people feeling isolated at parkrun as it is something that goes against the fundamental spirit of parkrun; for the event to be welcoming to all (parkrun Ireland, 2021b).

The finding in this study which has global implications is that parkrun can be considered as a community initiative, rather than a sport. The parkrun model could therefore be used with other physical activities to include a purposeful activity with social connection and an option to volunteer. This type of activity has potential to improve social and mental wellbeing in local communities.

### Methodological considerations

One methodological consideration is that the survey questions did not measure the quality of the social interactions. A brief verbal exchange or a lengthy conversation were both measured as one interaction, although they could have different impacts on the parkrun participant. Further consideration to the exact nature of the social interactions is recommended for future studies, perhaps by using interviews with parkrun participants to ask detailed questions about the type, length and value of each interaction and opportunity at parkrun.

Reporting results for each element of the social questions (Figure 1), rather than combing the results into a social score, increases the risk of a Type I error (Field, 2018). As the social questions were not designed to be reported as a combined score this cannot be avoided but should be considered when interpreting the results.

The data for this study was collected before the global COVID-19 pandemic. All parkrun events were paused during the pandemic in 2020 and have returned since 2021 (parkrun Ireland, 2021a). There is a growing body of research exploring the value that people have on social interaction since the pandemic and the impact of this on communities and physical activity (Bian, 2020; Bowe et al., 2022; Landmann & Rohmann, 2022; Stevenson, Wakefield, Felsner, Drury, & Costa, 2021). Care should be taken in interpreting and applying the results of this study in a post-pandemic environment, taking these factors into account.

The parkrun Health and Wellbeing Survey was only available in English language. The official first language of Ireland is Irish (Office of the Attorney General, 2020) with 4.2% of the population speaking Irish daily outside of the education system (Central Statistics Office, 2017). Any future surveys should be offered in both languages to include the Irish speaking population.

The social questions were designed specifically for the parkrun population. However, this means that the questions are not validated and direct comparisons with other populations or community groups cannot be made. As the survey respondents represented only a sample of parkrun participants from Ireland care should be taken when extrapolating the results to the whole parkrun population.

## Conclusions

This study considered the social interactions and opportunities of parkrun participants in Ireland. Over two thirds of survey respondents interacted with other parkrun participants and felt participation made them feel part of a community. Similar proportions interacted with strangers as with friends and with people from different and similar backgrounds which suggests a balance between opportunities for bonding and bridging social capital at parkrun. Volunteering at parkrun increased the potential for social interactions and opportunities.

Viewing parkrun as a community initiative rather than just a sport or physical activity allows for an appreciation of the social interactions and opportunities for participants which may support mental wellbeing. Volunteering at parkrun events increases the potential for this benefit of parkrun participation. The parkrun model could be applied to new community physical activity initiatives to promote social and mental wellbeing. Future research could explore the depth and extent of social interactions at parkrun, and why certain people feel isolated at parkrun events and similar community initiatives. This has implications for the design and implementation of new community initiatives which are a low cost, yet impactful, method of supporting the mental and social wellbeing of the population.

## Data Availability

All data produced in the present work are contained in the manuscript.

## Acknowledgements

The authors would like to thank all the parkrun participants who completed the survey and to thank the parkrun Research Board for their support and guidance. Thank you to the Advanced Wellbeing Research Centre Public Involvement in Research Group for their assistance in writing a lay summary. We would also like to thank Dr Gareth Wiltshire, Loughborough University, for his advice on the sociological concepts contained in this report. Finally, we would like to thank Dr Lisa Pursell, University of Galway, for her input relating to the statistical analysis of the data.

## Author contributions

AB, HQ and SH designed the survey, sampling method and analysis plan; AD and SH were responsible for the statistical analysis. AD, HQ, AB, and SH drafted the manuscript. All authors contributed to the writing of and approval of the final manuscript.

## Open access declaration

For the purpose of open access, the author has applied a Creative Commons Attribution (CC BY) licence to any Author Accepted Manuscript version arising from this submission.

## Ethics approval and consent to participate

The research design and consent procedures for the Health and Wellbeing Survey were reviewed and approved by Sheffield Hallam University Research Ethics Committee (Reference number: ER7034346). Written informed consent was received from all participants via the first page of the online survey. The research design for this secondary data analysis was approved by Sheffield Hallam University Research Ethics Committee (Reference number: ER23166109).

## Supplementary material

**Supplementary Table S1.** Proportion of respondents, by gender, who selected each response to social questions.

|  | n |  |  | % |  |  | Compare male vs female |  |  |
| --- | --- | --- | --- | --- | --- | --- | --- | --- | --- |
| | Total | Male | Female | Total | Male | Female | $\chi^2$ (df=1) | p | Cramer's V |
| **Social interactions** |  |  |  |  |  |  |  |  |  |
| All respondents to this question | 3,946 | 1,453 | 1,713 | 100 | 45.9 | 54.1 |  |  |  |
| parkrun participants | 2,740 | 1,061 | 1,218 | 69.4 | 73.0 | 71.1 | 1.43 | 0.231 | 0.021 |
| friends | 1,812 | 682 | 810 | 45.9 | 46.9 | 47.3 | 0.04 | 0.845 | 0.003 |
| strangers | 1,740 | 703 | 754 | 44.1 | 48.4 | 44.0 | 6.03 | 0.014 | 0.044 |
| family | 1,171 | 415 | 552 | 29.7 | 28.6 | 32.2 | 4.97 | 0.026 | 0.040 |
| spouse/partner | 989 | 374 | 423 | 25.1 | 25.7 | 24.7 | 0.46 | 0.499 | 0.012 |
| member sports club | 829 | 310 | 372 | 21.0 | 21.3 | 21.7 | 0.07 | 0.795 | 0.005 |
| neighbours | 699 | 293 | 301 | 17.7 | 20.2 | 17.6 | 3.47 | 0.062 | 0.033 |
| work colleagues | 398 | 167 | 159 | 10.1 | 11.5 | 9.3 | 4.16 | 0.041 | 0.036 |
| members non sports club | 357 | 145 | 152 | 9.0 | 10.0 | 8.9 | 1.13 | 0.287 | 0.019 |
| did not interact | 231 | 89 | 88 | 5.9 | 6.1 | 5.1 | 1.45 | 0.228 | 0.021 |
| other | 85 | 26 | 43 | 2.2 | 1.8 | 2.5 | 1.92 | 0.166 | 0.025 |
| **Social opportunities** |  |  |  |  |  |  |  |  |  |
| All respondents to this question | 3,867 | 1,428 | 1,683 | 100 | 45.9 | 54.1 |  |  |  |
| I feel part of my community | 1,862 | 704 | 863 | 48.2 | 49.3 | 51.3 | 1.21 | 0.272 | 0.020 |
| I have met new people of a different background | 1,302 | 537 | 596 | 33.7 | 37.6 | 35.4 | 1.60 | 0.205 | 0.023 |
| I have met new people of a similar background | 1,253 | 500 | 566 | 32.4 | 35.0 | 33.6 | 0.66 | 0.418 | 0.015 |
| I feel closer to my existing friends of family | 871 | 304 | 396 | 22.5 | 21.3 | 23.5 | 2.23 | 0.136 | 0.027 |
| It's made no difference to me | 787 | 304 | 289 | 20.4 | 21.3 | 17.2 | 8.49 | **0.004** | 0.052 |
| It's made me feel isolated | 717 | 220 | 351 | 18.5 | 15.4 | 20.9 | 15.31 | **<0.001** | 0.070 |
| I have joined a sports club | 460 | 158 | 230 | 11.9 | 11.1 | 13.7 | 4.79 | 0.029 | 0.039 |
| I have joined a non-sports club | 97 | 28 | 55 | 2.5 | 2.0 | 3.3 | 5.08 | 0.024 | 0.040 |
| other | 97 | 34 | 50 | 2.5 | 2.4 | 3.0 | 1.02 | 0.312 | 0.018 |
| It has allowed me to spend time on my own | 0 | 0 | 0 | 0.0 | n/a | n/a | n/a | n/a | n/a |

$p < 0.01$ in bold male vs female.

**Supplementary Table S2.**
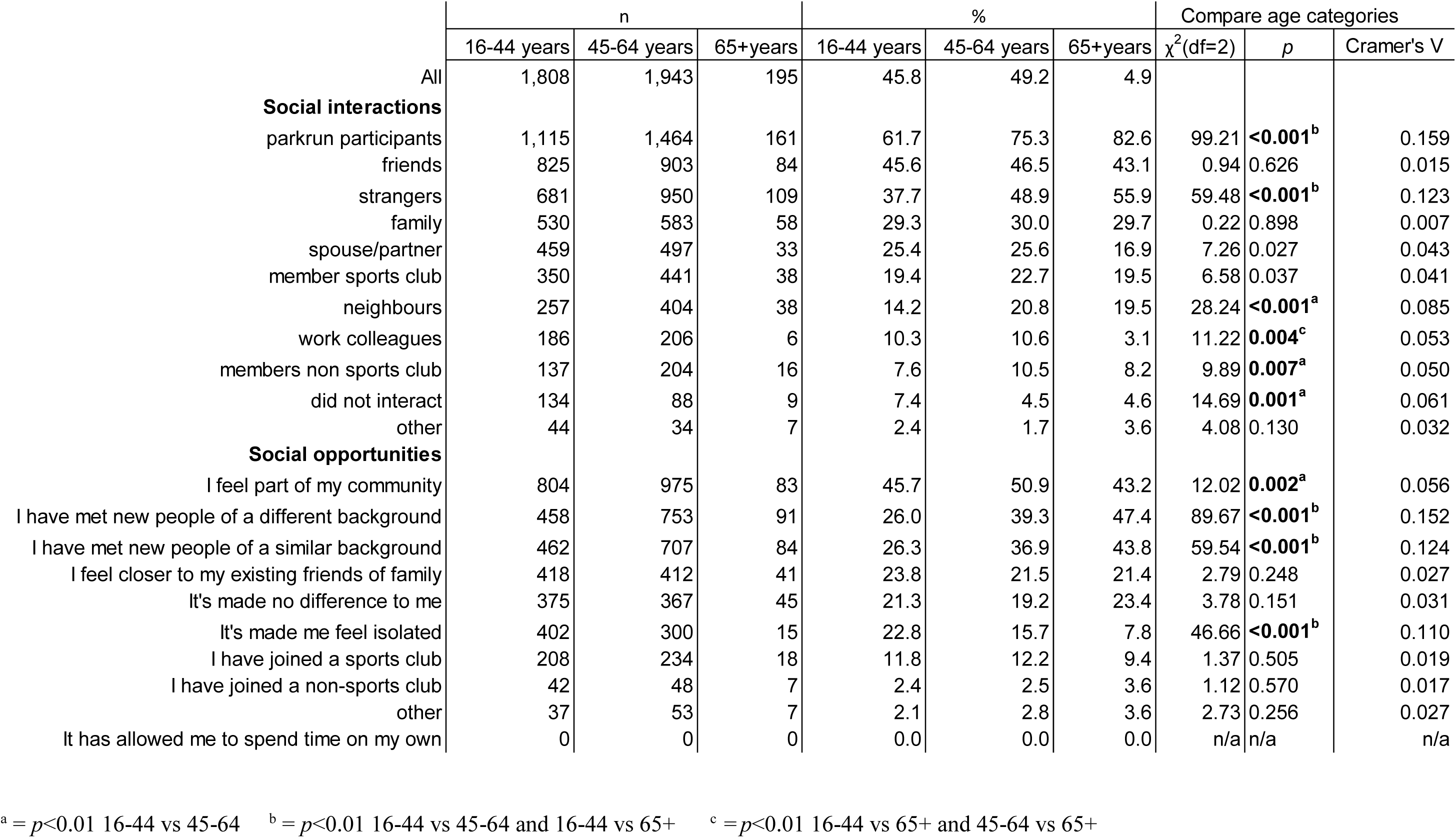
Proportion of respondents, by age category, who selected each response to social questions.

**Supplementary Table S3.** Proportion of respondents, by parkrun participation type, who selected each response to social questions.

|  | n |  |  | % |  |  | Compare participation types |  |  |
| --- | --- | --- | --- | --- | --- | --- | --- | --- | --- |
| | Runner/<br>walker and<br>volunteer | Runner/<br>walker only | Volunteer<br>only | Runner/<br>walker and<br>volunteer | Runner/<br>walker only | Volunteer<br>only | $\chi^2$ (df=2) | p | Cramer's V |
| All | 1,805 | 2,095 | 46 | 45.7 | 53.1 | 1.2 |  |  |  |
| **Social interactions** |  |  |  |  |  |  |  |  |  |
| parkrun participants | 1,563 | 1,140 | 37 | 86.6 | 54.4 | 80.4 | 475.72 | <0.001<sup>b</sup> | 0.347 |
| friends | 999 | 792 | 21 | 55.3 | 37.8 | 45.7 | 120.15 | <0.001<sup>a</sup> | 0.174 |
| strangers | 1,063 | 654 | 23 | 58.9 | 31.2 | 50.0 | 301.91 | <0.001<sup>a</sup> | 0.277 |
| family | 602 | 550 | 19 | 33.4 | 26.3 | 41.3 | 26.43 | <0.001<sup>a</sup> | 0.082 |
| spouse/partner | 535 | 441 | 13 | 29.6 | 21.1 | 28.3 | 38.35 | <0.001<sup>a</sup> | 0.099 |
| member sports club | 534 | 289 | 6 | 29.6 | 13.8 | 13.0 | 147.45 | <0.001<sup>a</sup> | 0.193 |
| neighbours | 456 | 234 | 9 | 25.3 | 11.2 | 19.6 | 132.24 | <0.001<sup>a</sup> | 0.183 |
| work colleagues | 220 | 176 | 2 | 12.2 | 8.4 | 4.3 | 17.03 | <0.001<sup>a</sup> | 0.066 |
| members non sports club | 250 | 101 | 6 | 13.9 | 4.8 | 13.0 | 96.97 | <0.001<sup>a</sup> | 0.157 |
| did not interact | 28 | 203 | 0 | 1.6 | 9.7 | 0.0 | 119.42 | <0.001<sup>a</sup> | 0.174 |
| other | 31 | 51 | 3 | 1.7 | 2.4 | 6.5 | 6.58 | 0.037 | 0.041 |
| **Social opportunities** |  |  |  |  |  |  |  |  |  |
| I feel part of my community | 1,137 | 692 | 33 | 63.4 | 34.1 | 68.8 | 337.00 | <0.001<sup>b</sup> | 0.295 |
| I have met new people of a different background | 944 | 335 | 23 | 52.7 | 16.5 | 47.9 | 561.95 | <0.001<sup>b</sup> | 0.381 |
| I have met new people of a similar background | 868 | 367 | 18 | 48.4 | 18.1 | 37.5 | 400.60 | <0.001<sup>b</sup> | 0.322 |
| I feel closer to my existing friends of family | 421 | 443 | 7 | 23.5 | 21.8 | 14.6 | 3.07 | 0.215 | 0.028 |
| It's made no difference to me | 184 | 600 | 3 | 10.3 | 29.6 | 6.3 | 224.75 | <0.001<sup>b</sup> | 0.241 |
| It's made me feel isolated | 281 | 432 | 4 | 15.7 | 21.3 | 8.3 | 23.07 | <0.001<sup>a</sup> | 0.077 |
| I have joined a sports club | 305 | 151 | 4 | 17.0 | 7.4 | 8.3 | 83.73 | <0.001<sup>a</sup> | 0.147 |
| I have joined a non-sports club | 57 | 37 | 3 | 3.2 | 1.8 | 6.3 | 10.08 | 0.006<sup>b</sup> | 0.051 |
| other | 40 | 57 | 0 | 2.2 | 2.8 | 0.0 | 2.526 | 0.283 | 0.026 |
| It has allowed me to spend time on my own | 0 | 0 | 0 | 0.0 | 0.0 | 0.0 | n/a | n/a | n/a |

<sup>a</sup> = $p < 0.01$ runner/walker and volunteer vs runner/walker only<sup>b</sup> = $p < 0.01$ runner/walker and volunteer vs runner/walker only and volunteer only vs runner/walker only

